# Predictors of Perioperative Mortality in Paediatric Surgery at a Tertiary Hospital in Sagamu, Nigeria

**DOI:** 10.1101/2025.10.14.25337967

**Authors:** Lukmon O. Amosu, Chigbundu C. Nwokoro, Oluwakemi A. Shotayo, Ibukunolu O. Ogundele, Adeleke O. Adekoya, Adekunle O. Ajayi, Solomon O. Ariyibi, Adeduntan S. Olagbenro, Olusola A. Sogebi, Lateef O.A. Thanni

## Abstract

**Background:** The burden of surgical diseases in children and perioperative complications in sub-Saharan Africa remains high. The assessment of the pattern and the determinants of perioperative mortality plays an invaluable role in identifying indicators of poor outcomes in order to improve overall outcomes in children’s surgery, thus necessitating this study.

**Methods:** A retrospective review of medical records of patients aged 15 years and below, who had general pediatric, oncological, and urological surgeries under general anesthesia between January 2014 and December 2023, with complete records up to at least post-surgery or died within 30 days of surgery, was carried out. Information extracted included biodata, diagnosis, ASA classification of physical status, time of death after surgery, cause of death, and duration of surgery. Data were collated and analyzed using univariate and multivariate statistical tools.

**Results:** A total of 1621 patients were analyzed. The 30-day perioperative mortality rate in this study was 2.96% (296 per 10,000 patients); jejuno-ileal atresia, gastrochisis and bladder exstrophy are the conditions associated with the highest mortality rates, well above 50%. Furthermore, logistic regression model identified neonatal age group, ASA class greater than II and repeated surgical procedures as the significant predictors of mortality, while sepsis and intestinal/anastomosis failure were identified as the most common direct cause of death.

**Conclusion:** Neonatal age group, ASA class greater than II, and repeated surgery are the significant predictors of mortality in children’s surgery in our practice. Efforts should be made to combat sepsis and provide physiologic support and intensive care to improve outcomes.

**Key messages:** What is already known about this subject:

- Perioperative mortality rate in pediatric surgery in sub-Saharan Africa remains high.
- Only a few studies in the region have attempted to identify predictors of POM in children.

**What this study adds:**

- This study revealed the real burden of POM in our practice and identified the determinants of poor outcomes
- Neonatal age group, high ASA classification and repeated surgeries are significant predictors of mortality in this study
- Jejuno-ileal atresia and gastrochisis are associated with a high case fatality rate.

**How this study might affect research, practice or policy:**

- Findings from this study provide potential tools to improve outcomes in pediatric surgery in the region.

## INTRODUCTION

Sub-Saharan Africa is a region that has been associated with a high incidence of perioperative mortality in children, especially neonates and infants. ^1,2^ Even though access to pediatric surgical services has improved significantly in the region in the new millennium, and in spite of the overall gains in global surgery and collaboration, the burden of surgical diseases in children and perioperative complications, including death in low- and middle-income countries (LMICS), remains high.^3,4^ Thus, there is still a need for continuous attention to the outcomes of surgery and indicators of poor surgical outcomes in the region. Authorities have advocated for the inclusion of perioperative mortality as a health indicator of the quality and safety of surgery and anesthesia.^5^

It is worth noting that despite the displeasing mortality reports coming from sub-Saharan Africa, surgical outcomes in the region are still largely underreported.^5^ Perioperative mortality (POM) rate is defined as the number of deaths per population on the day of surgery and before discharge from the hospital or within 30 days of surgery, whichever is earlier, or the percentage of perioperative deaths per total operated cases in a given period.^5,6^ Some researchers have reported POM rates of 156 per 10,000 procedures (1.56%), 170 per 10,000 procedures (1.7%), and 1.71% in Nigeria, Ghana and Kenya respectively.^4,7,8^ A higher POM rate of 2.54% was, however, reported from Ethiopia.^6^

The assessment of the rate and pattern of POM plays an invaluable role in identifying indicators of poor outcomes and will assist in formulating policies to improve overall outcomes in children’s surgery and anesthesia. More so, risk-adjusted outcomes data are needed to evaluate surgical quality and for proper planning, as well as the distribution of resources and expert staffing for the higher-risk children ^3,7^ A previous study conducted 13 years ago in this hospital on mortality rate and patterns did not identify determinants of mortality, and was not specifically focused on children.^9^ Children are a particularly vulnerable group, and they determine the future and progress of any nation. It is thus expedient that issues relating to their survival and well-being be given utmost attention and regard.

Globally, congenital anomalies and prematurity are common factors associated with mortality in children’s surgery.^10^ However, the few regional data available often reveal neonatal/ infant age group, emergency surgeries being the main predictors of mortality, while sepsis is frequently the most common direct cause of death.^6,7^ It is imperative to understand the etiology and pattern of mortality in our local setting and analyze the determinants of fatal outcomes in our pediatric surgical practice. Thus, this study was undertaken to determine the burden of POM and to assess the determinants of mortality in children who had surgery under general anesthesia at the hospital.

## METHODOLOGY

### Settings

This research work was carried out at the Department of Surgery of the Olabisi Onabanjo University Teaching Hospital, Sagamu, Ogun State (OOUTH). The hospital is a 420-bed tertiary referral hospital situated in a semi-urban area of Southwestern Nigeria. There is a pediatric surgery unit and a dedicated 12-bed pediatric surgery ward, with an additional number of bed spaces in the general children’s ward. Surgical neonates are nursed in the 32-bed general neonatal ward alongside other medical cases. Ethics approval to conduct this study was obtained from the hospital’s Health Research Ethics Committee with approval number OOUTH/HREC/011/012/E118/2024AP.

### Study design and study population

This study is a clinical retrospective, analytical study that involved the review of the medical records of patients who had surgery between January 2014 and December 2023, including the review of theatre operation records, central mortality records and patients’ case files. The study employed a total population sampling technique in order to capture all eligible patients and have a true picture of the mortality rate. Inclusion criteria for the study were all patients aged 15 years and below who had general pediatric, oncological and urological surgeries under general anesthesia with complete records up to at least 30days post-surgery or died within 30 days of surgery. Patients whose available data are grossly inadequate for analysis were excluded from the study.

### Data collection

The records files of all the patients that met the inclusion criteria were thoroughly reviewed, extracting information on biodata, clinical diagnosis/indication for surgery, nature of surgery (elective versus emergency), American Society of Anesthesiology (ASA) classification of physical status, time of death after surgery, cause of death, anesthesia technique, duration of surgery and anesthesia using a structured checklist. The time of death postoperatively was classified into three categories, viz: first 24 hours (immediate mortality), within 7 days (early mortality), 8 days to 30 days (late mortality). The information retrieved generated the data, which was entered into a spreadsheet, cleaned and collated appropriately.

### Data analysis

Data were analyzed using the Statistical Package for Social Sciences (SPSS) version 26 (IBM Corp., Armonk, NY, USA). The results are presented as tables and graphs. The mortality rates were calculated as the total number of deaths divided by the total number of operated patients, expressed in ratios per 10,000 patients, as well as in percentages. All variables potentially associated with 30-day perioperative mortality were subjected to Chi-square analysis or Fisher’s exact test. Variables with significant association with perioperative mortality were further analyzed using a multivariate regression model to determine the predictor(s) of mortality. A p-value of less than 0.05 was considered statistically significant.

## RESULTS

A total of 1706 children had surgical operations during the period under review; however, 1621 patients had adequate data in the available records for review. The age range of the patients was between 1 day and 15 years, with a median age of 3 years. Out of the 1621 patients, 1270 were males and 351 were females, with a Male: Female ratio of 3.6:1. A total of 1065 patients (65.7%) had their surgery as elective cases, while the rest were performed as emergency procedures. The majority of the patients belong to ASA class 1 (65.4%). Table 1 shows the details of the clinical and demographic characteristics of the patients.

**Table 1:** Clinical and demographic characteristics of patients and mortality rates.

| Parameter |  | Number of patients (%) | 30-day Peri-operative Death (n) | Mortality rate per category (%) | P-value* |
| --- | --- | --- | --- | --- | --- |
| Total |  | 1621 | 48 | 2.96 |  |
| Age group | < 1 month | 114 (7.0) | 35 | 30.7 | < 0.001 |
|  | 1 – 11 months | 303 (18.7) | 8 | 2.6 |  |
|  | 1 – 5 years | 541 (33.4) | 2 | 0.3 |  |
|  | 6 – 10 years | 379 (23.4) | 1 | 0.3 |  |
|  | 11 – 15 years | 284 (17.5) | 2 | 0.7 |  |
| Sex | Male | 1270 (78.3) | 38 | 2.9 | 0.89 |
|  | Female | 351 (21.7) | 10 | 2.8 |  |
| ASA classification | I | 1060 (65.4) | 2 | 0.2 | <0.001 |
|  | II | 422 (26.0) | 18 | 4.3 |  |
|  | III | 136 (8.4) | 26 | 19.1 |  |
|  | IV | 3 (0.2) | 2 | 66.7 |  |
| Surgery urgency | Emergency | 556 (34.3) | 40 | 7.2 | <0.001 |
|  | Elective | 1065 (65.7) | 8 | 0.8 |  |
| Number of procedures per patient | Single procedure | 1593 (98.3) | 39 | 2.4 | <0.001 |
|  | More than one procedure | 28 (1.7) | 9 | 32.1 |  |

\* = Chi-square test comparing category with surgical outcome

The 24-hour perioperative mortality rate in this study was 0.87% (87 per 10000 patients), while the total 30-day perioperative mortality rate was 2.96% (296 per 10,000 patients). Also, as shown in Table 1, further analysis revealed that neonates have the highest mortality rate of 30.5%; the mortality rate decreased significantly with increasing age (P-value <0.001). However, for ASA classification of physical status, the mortality rate increased significantly with increasing ASA class (P-value <0.001). There was also a significantly higher mortality rate in emergency procedures compared to elective surgical procedures (7.2% vs 0.8%), as well as a higher mortality rate in patients who had repeated surgeries compared with single surgery (32.1% vs 2.4%). No significant difference was observed in the mortality rate between males and females.

In order to relate mortality with primary surgical pathology in these patients, case fatality rates were computed for all surgical conditions that resulted in mortality and are presented in Table II. Case fatality review revealed jejuno-ileal atresia, gastrochisis, and bladder exstrophy as conditions with very high fatality rates in this review, with case fatality rates well above 50%. Other conditions, such as omphalocele, malrotation/ volvulus, enterocutaneous fistula and duodenal atresia, also have significantly high fatality rates in the range of 21% to 33%.

**Table II.** Case fatality rate.

| Surgical pathology | Total number<br>of cases | 30-Day peri-<br>operative death | Mortality<br>rate (%) |
| --- | --- | --- | --- |
| Omphalocele | 14 | 3 | 21.4 |
| Gastrochisis | 6 | 4 | 66.7 |
| Jejuno-ileal atresia | 14 | 8 | 57.1 |
| Malrotation/volvulus | 19 | 5 | 26.3 |
| Anorectal malformation | 81 | 6 | 7.4 |
| Duodenal atresia | 16 | 4 | 25.0 |
| Intussusception | 123 | 3 | 2.4 |
| Obstructed/ strangulated hernia | 44 | 2 | 4.5 |
| Hirschsprung's disease | 67 | 3 | 4.5 |
| Enterocutaneous fistula | 3 | 1 | 33.3 |
| Wilm's tumor | 12 | 1 | 8.3 |
| Biliary atresia | 8 | 2 | 25.0 |
| Bladder exstrophy | 2 | 2 | 100 |
| Cleft lip/ palate | 5 | 1 | 20.0 |
| Pelvi-ureteric Junction obstruction | 11 | 1 | 9.1 |
| Esophageal stricture | 3 | 1 | 33.3 |

Furthermore, in identifying possible predictors of mortality in these patients, a binary logistic regression analysis was done, which identified neonatal age group, ASA class greater than II, and repeated surgical procedures as risk factors for mortality. Although surgical urgency (emergency vs elective) showed a significant effect in univariate analysis, it lost significance in the multivariate analysis. The finding is presented in Table III. Further analysis also identified the specific cause of death in the 48 cases of mortality. Sepsis/ Multiple Organ Dysfunction Syndrome (MODS) was the most common immediate cause of death, accounting for 35.5%, followed closely by intestinal/anastomosis failure, accounting for 20.8%. Other identified immediate causes of death include respiratory failure (12.5%), fluid overload (8.3%), reactionary hemorrhage (8.3%), and aspiration pneumonitis 4.2%.

**Table III:** Binary regression analysis of predictors of mortality.

| Variables |  | Adjusted<br>Odd Ratio | 95%<br>confidence<br>interval | P-value |
| --- | --- | --- | --- | --- |
| **Univariate**<br>**analysis** |  |  |  |  |
| Age | < 1 month<br>1month -15yrs (ref) | 50.92 | 25.91 –100.06 | <0.001 |
| ASA classification | ASA I -II (ref)<br>ASA III-IV | 18.44 | 10.07 –33.78 | <0.001 |
| Surgery urgency | Elective (ref)<br>Emergency | 0.10 | 0.05 –0.21 | <0.001 |
| Repeat surgical<br>procedure | Single procedure<br>(ref)<br>More than one<br>procedure | 0.05 | 0.02 –0.13 | <0.001 |
| **Multivariate**<br>**analysis** |  |  |  |  |
| Age | < 1 month<br>1month -15yrs (ref) | 55.58 | 21.66 – 142.63 | <0.001 |
| ASA classification | ASA I -II (ref)<br>ASA III-IV | 15.42 | 6.56 – 36.28 | <0.001 |
| Surgery urgency | Elective (ref)<br>Emergency | 0.65 | 0.22 – 1.96 | 0.45 |
| Repeat surgical<br>procedure | Single procedure<br>(ref)<br>More than one<br>procedure | 8.70 | 2.67 – 28.38 | <0.001 |

The postoperative period was also analyzed based on the period of occurrence of death. As shown in Figure 1, in the immediate postoperative period (first 24 hours postoperative), sepsis, respiratory failure, reactionary hemorrhage, and anesthesia adverse events are the most common causes of death. However, in the early postoperative period (one to seven days postoperative), sepsis, fluid overload and respiratory failure are the most common causes. Late postoperative deaths (later than seven days postoperative) are mostly due to anastomosis/ intestinal failure and sepsis.

**Figure 1:**
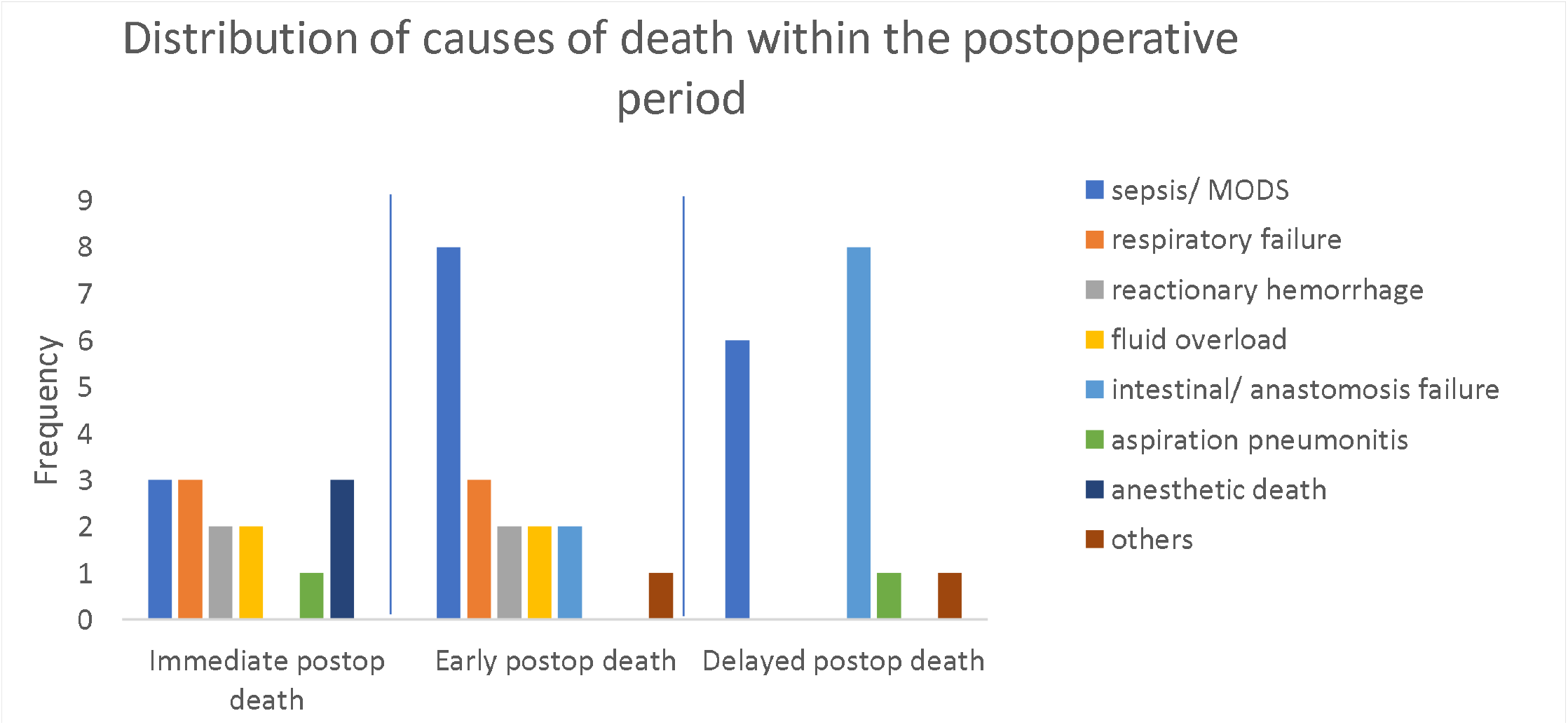
Showings the frequncy distribution of the causes of death within the post operative period. Sepsis is the most common cause of death featuring in all the three peroid categories.

## DISCUSSION

This study provided the much-needed attention to pediatric surgical outcomes in a sub-Saharan African setting, focusing on perioperative mortality rate and factors associated with mortality in pediatric surgery, as well as developing possible models to predict perioperative mortality in the age group. The result showed good distribution of patients across the different age groups within the pediatric population. Although the majority of the patients reviewed were male, this is simply a reflection of male-predominant pathologies in the cases reviewed.

The overall 30-day mortality rate in this study is reported as 2.96%, which is similar to the findings of Talabi et al^11^ in Nigeria and Tarekegn et al^6^ in Ethiopia (3.2% and 2.54% respectively), and within the global range of 0.3% to 3.6 % reported by Tangel et al.^12^ However, most studies from high-income countries reported rates less than 1%.^13^ A multinational study by the global health collaborative group^14^ described the disparity between the mortality rates among high-, middle- and low-income countries in emergency abdominal surgery, where 30-day mortality rates were found to be 4 to 7 times higher in middle and low-income countries compared to high-income countries. Our finding may be a reflection of the sub-Saharan African situation, and it provides a deep insight into the quality and safety of surgical and anesthesia services in our pediatric surgical practice.

In this study, factors identified to be associated with perioperative mortality included neonatal age, high ASA classification status, emergency surgery, and multiple surgeries within a single admission. This finding is quite similar to earlier reports from low- and middle-income as well as high-income countries.^6,10,11^ Other commonly reported factors, such as prematurity and the presence of congenital anomalies, were not reviewed in this study due to inadequate data as a result of the retrospective nature of the study. A younger age is the most important predictor of mortality in this study; the mortality rates in neonates are quite high (30.7%), with neonates having about 55-fold increased odds and a higher risk of perioperative death compared to older children (see Table III). This appears to be a consistent finding in sub-Saharan Africa, as other authors in the region had reported similar alarming rates.^7,15^ ASA class greater than II is also associated with about 15-fold increased odds and a higher risk of perioperative mortality compared to lower ASA class status. The above findings underscore the challenges of neonatal and high-risk surgeries in our setting. These categories of patients often require high-end physiologic support and intensive care, which are mostly lacking or inadequate in our setting. Although emergency surgery showed a significant effect on the mortality rate when compared to elective surgeries in the univariate analysis, it became insignificant in the multivariate analysis. This may suggest that the effect earlier noticed could be accounted for by other strong factors, such as age and ASA classifications. Expectedly, the performance of repeated surgery during a single admission is associated with a slight increase in risk of mortality, as such patients are those that had developed a form of postoperative complication, predisposing them to poorer outcomes. Talabi et al^7^ also reported a similar finding. However, it would have been more informative to classify the postoperative complications suffered by these patients and correlate them with the mortality rate, but this is not feasible due to the retrospective nature of this study, as available data were insufficient.

This study also looked into the direct etiology of deaths in the perioperative period. Sepsis, intestinal/anastomosis failure and respiratory failure are the more common specific causes of death. This is also similar to the findings from other low- and middle-income countries.^2,7^ However, in a report by de Bruin et al^10^ from a high-income country, congenital cardiac anomalies were identified as the most common cause of death, although sepsis also had a significant contribution. Our study is unable to evaluate associated congenital anomalies due to inadequate data.

Furthermore, looking into case-specific mortality, gastrochisis, jejuno-ileal atresia, and midgut volvulus are among the cases mostly associated with mortality: these conditions are often associated with sepsis, intestinal failure and anastomotic challenges – the common etiology of death in this review. In addition, these challenges are prevalent in many reports from sub-Saharan Africa^11^ and represent regional setbacks in these aspects of the critical care of patients. Other factors, such as fluid overload and aspiration pneumonitis, also implicated in some cases of mortality, are reflections of poor nursing care and inadequate monitoring still plaguing our practice. It was also reported by earlier researchers in the region, but the prevalence appears to be reducing.^11^

The key strength of this study is its ability to reveal the burden of POM in our practice and also predict determinants of poor outcomes, as well as the use of empirical data to identify critical areas of challenges in our local setting. Based on the findings of this study, it could be inferred that to improve children’s survival after surgery, there is a need to pay more attention to determinants of mortality, such as neonatal surgery, surgery in critically ill children, and emergency surgery. Thus, improving capacity in neonatal intensive care, peri-operative ventilation support, adequate monitoring, nutritional support, including parenteral nutrition, are among the important interventions for better outcomes. In addition, it is apparent from Figure 1 that the immediate challenges to a postoperative patient in our practice are respiratory failure, anesthesia adverse events and fluid overload; these could be mitigated by providing intensive care and adequate monitoring. The major late postoperative challenges are sepsis and intestinal and anastomosis failure; consequently, there is a need for concerted effort towards strategies to improve nutritional support and prevent and treat sepsis in the perioperative period.

### Limitation

The retrospective nature of this study limited the array of predictive variables that could be tested due to insufficient data. A prospective follow-up study exploring all possible variables is necessary. In conclusion, the 30-day perioperative mortality in this study is 2.96%. Neonatal age group, ASA class greater than II, and repeated surgery within a single admission are significant predictors of mortality in children’s surgery in our practice.

## Data Availability

All data produced in the present study are available upon reasonable request to the authors.

## Conflict of interest

none to declare

## Funding

The authors received no funding or grants from any organization related to the study.

## Notes

### Competing Interest Statement

The authors have declared no competing interest.

### Funding Statement

This study did not receive any funding

### Author Declarations

Ethics committee/IRB of Olabisi Onabanjo University Teaching Hospital, Sagamu, Ogun State, Nigeria gave ethical approval for this work.

